# Naloxone Administration and Survival in Overdoses Involving Opioids and Stimulants: An Analysis of Law Enforcement Data from 63 Pennsylvania Counties

**DOI:** 10.1101/2024.08.27.24312661

**Authors:** Manuel Cano, Abenaa Jones, Sydney M. Silverstein, Raminta Daniulaityte, Frank LoVecchio

## Abstract

**Background:** In consideration of rising opioid-stimulant deaths in the United States, this study explored rates of naloxone administration and survival in suspected opioid overdoses with, versus without, stimulants co-involved.

**Methods:** The study analyzed 26,635 suspected opioid-involved overdoses recorded by law enforcement/first-responders in the Pennsylvania Overdose Information Network in 63 Pennsylvania counties, January 2018-July 2024. All measures, including suspected drug involvement, were based on first-responder assessment/report. Relative frequencies and chi-square tests were first used to compare suspected opioid overdoses with, versus without, stimulants (cocaine or methamphetamine) co-involved. Next, mediation analyses tested naloxone administration as a mediator in the association between stimulant co-involvement (in opioid overdoses) and survival.

**Results:** Naloxone was reportedly administered in 72.2% of the suspected opioid-no-cocaine overdoses, compared to 55.1% of the opioid-cocaine-involved overdoses, and 72.1% of the opioid-no-methamphetamine overdoses vs. 52.4% of the opioid-methamphetamine-involved overdoses. With respect to survival rates, 18.0% of the suspected opioid-no-cocaine overdoses ended in death, compared to 41.3% of the opioid-cocaine overdoses; 18.1% of the opioid-no-methamphetamine overdoses ended in death, versus 42.9% of the opioid-methamphetamine overdoses. In mediation analyses (adjusted for demographics, county, year, and other drug co-involvement), naloxone administration mediated 38.7% (95% Confidence Interval [CI], 31.3%-46.0%) of the association between suspected cocaine co-involvement and survival and 39.2% (95% CI, 31.3%-47.1%) of the association between suspected methamphetamine co-involvement and survival.

**Conclusions:** Among suspected opioid overdoses recorded in the Pennsylvania Overdose Information Network, stimulant co-involvement was associated with lower naloxone administration and higher fatality, with naloxone partially mediating the association between stimulant co-involvement and death.

## Introduction

The current phase (“fourth wave”) of the United States’ (US) overdose crisis is characterized by rising numbers of overdoses involving synthetic opioids (e.g., non-pharmaceutical fentanyl) and stimulants (e.g., cocaine, methamphetamine) (Ciccarone, 2021). Rising opioid-stimulant co-use or co-exposure in the US has been documented across data from emergency department visits, postmortem toxicology, drug seizures, substance use treatment admission records, case reports, and population-based or community-based surveys (Jones et al., 2020). Research with individuals who use drugs has identified diverse opioid-stimulant co-use motivations, including: achieving synergistic effects; counteracting unfavorable effects of one drug type by also using another drug type; reducing perceived overdose risk; managing opioid withdrawal symptoms; and adapting to changes in the cost and availability of different drugs (Daniulaityte et al., 2022; Fredericksen et al., 2024; Silverstein et al., 2021; Valente et al., 2020).

Increasing opioid-stimulant co-use represents a growing public health concern for several reasons, including overdose risk; several studies have documented higher rates of non-fatal and fatal overdoses among individuals who co-use opioids and stimulants (relative to those who use opioids only; Karamouzian et al., 2024; Korthuis et al., 2022; Palis et al., 2022). Between 2015 and 2022, the proportion of US overdose deaths involving stimulants rose from 23% to 53% (Centers for Disease Control and Prevention, 2024a), and opioids were co-involved in 78.6% and 65.7% of overdose deaths involving cocaine and psychostimulants (e.g., methamphetamine), respectively (in 2021; Spencer et al., 2023).

In cases of opioid-stimulant overdose, timely naloxone administration and supportive care is recommended to counteract the effects of the opioids involved in the overdose (Ahmed et al., 2022; Britch & Walsh, 2022; Rzasa Lynn & Galinkin, 2018). Naloxone, an opioid overdose reversal medication, is used to restore breathing in cases of opioid-induced respiratory depression (Davis & Carr, 2020; Morris & Kleinman, 2020). Naloxone can be administered by medical personnel, nonmedical first responders, and bystanders, and because time is of the essence when responding to an opioid-involved overdose (Davis & Carr, 2020), increasing naloxone access for first responders and community members has been identified as a public health priority (Centers for Disease Control and Prevention, 2024b).

As opioid-related overdose deaths continue to increase nationally, there have been efforts to expand access to naloxone among law enforcement, who are often first responders at the scene of an overdose (Ray et al., 2015; Wagner et al., 2016). In Pennsylvania, the setting of the present study, legislation first enacted in 2014 (Act 139) enables police to administer naloxone and complete online training, and in a survey of 980 Pennsylvania police four years after the legislation’s implementation, 91% reported access to naloxone and 73% indicated that they were typically first on the scene of an overdose (Jacoby et al., 2020).

Opioid-stimulant-involved overdoses may be particularly complex and risky (Pergolizzi et al., 2021; Shastry et al., 2024), as these overdoses involve multiple substances, each with unique as well as potentially interactive effects (Shastry et al., 2024). A 2024 study of opioid overdose patients in nine US emergency departments found that cases of opioid-stimulant co-exposure required more doses of naloxone than opioid-only cases (Shastry et al., 2024). Moreover, research suggests that the signs/symptoms/clinical presentations of overdoses involving opioid-stimulant combinations can differ from those observed in cases of opioid-only or stimulant-only toxicity (Glidden et al., 2023), potentially contributing to first responders’ or bystanders’ difficulty identifying what drugs may be involved and how best to respond.

In a 2022 US survey, police chiefs, especially in the Northeast, expressed relatively less confidence in their officers’ readiness to respond to methamphetamine-involved overdoses versus opioid overdoses (Bailey et al., 2024). It is unclear to what extent first responders such as police tend to administer naloxone during overdose calls in which the use of both opioids and stimulants is suspected. In an analysis of fatal overdoses in Tennessee, naloxone had been reportedly administered in similar proportions (approximately 20%) of the overdoses involving opioids with or without stimulants, although information on naloxone administration was missing for nearly half of decedents (Korona-Bailey et al., 2021), and it is unknown how these figures might have differed if nonfatal (in addition to fatal) overdoses could have also been examined.

### The Present Study

In light of the complexity of opioid-stimulant-involved overdoses (Pergolizzi et al., 2021; Shastry et al., 2024) and recent increases in opioid-stimulant fatalities across the US (Ciccarone, 2021), this study examines naloxone administration and survival in overdoses involving opioids with or without stimulants (cocaine or methamphetamine). The study uses data from nonfatal and fatal overdose events recorded by law enforcement or other first responders in Pennsylvania’s Overdose Information Network (ODIN). The study investigates the following questions:

a. How do rates of naloxone administration and survival compare between suspected opioid-stimulant and opioid-no-stimulant overdoses recorded in ODIN?
b. To what extent might naloxone administration mediate the association between suspected stimulant co-involvement in opioid overdose and whether or not the individual survived the overdose event?

Overall, the study aims to fill gaps in the literature on rates of first responder naloxone administration and overdose fatality in opioid-stimulant overdoses; findings may help inform bystander and first responder training/protocols regarding responding to rising opioid-stimulant overdoses across the US.

Pennsylvania, the setting of the present study, is located in the northeastern US and has the fifth highest number of drug overdose deaths of any state nationwide (according to 2022 data; CDC, 2024a). Pennsylvania is divided into 67 counties, with the highest population in Philadelphia County (1.6 million), followed by Allegheny County (1.2 million, location of Pittsburgh) (CDC, 2024a). Nineteen of Pennsylvania’s counties are classified as urban, and 48 as rural; 26% of the state population resides in a rural county (Center for Rural Pennsylvania, n.d.).

Pennsylvania’s Overdose Information Network (ODIN) is the source of the data used in this study. ODIN was developed in 2018 as a collaboration between the Pennsylvania State Police and the Liberty Mid-Atlantic High Intensity Drug Trafficking Area, with the aim of offering “real-time information to aid in drug and overdose investigations and give leadership and policy makers reliable, comprehensive information in order to make informed decisions,” collecting data on “fatal and non-fatal drug overdoses, naloxone administrations and identifying markings found on drug packaging” (Rhoads, 2019, p. 2). In 2022, Pennsylvania legislation codified ODIN goals to include mapping and sending “overdose spike alerts” in cooperation with the Department of Health, also mandating that overdose incidents be entered by law enforcement within 72 hours (S.B. 1152, 2020; Lieberman & Davis, 2023).

ODIN data have been used in multiple peer-reviewed publications, including studies on racial/ethnic differences in overdose survival, naloxone, and drugs suspected in overdose (Barboza & Angulski, 2020), disparities in naloxone administration and survival in suspected opioid overdoses (Holmes et al., 2022), county differences in suspected synthetic opioid and heroin overdoses (Holmes & King, 2023), and suspected opioid overdoses before and after COVID-19 stay-at-home orders (King et al., 2021). ODIN is similar to the ODMAP Overdose Detection Mapping Application Program (High Intensity Drug Trafficking Areas Overdose Response Strategy, 2019) which has also been used in peer-reviewed publications on drug overdoses (Burgess-Hull et al., 2022; Piza et al., 2023). Although ODIN records lack the type of toxicological or medical information that informs data sources such as death certificates or emergency department admission records, ODIN represents one of the few data sources with information about naloxone administration at the scene of overdose, as well as both fatal and nonfatal overdoses, even some nonfatal overdoses in which the victim was not evaluated at the emergency department or transported by emergency medical services.

## Methods

### Data Source

The [blinded for peer review] Institutional Review Board designated the present study as exempt based on Federal Regulations 45CFR46(4). All data were drawn from the Pennsylvania State Police’s publicly available version of the Overdose Information Network dataset (Pennsylvania State Police, 2024) accessible via the Pennsylvania government’s OpenData website (https://data.pa.gov/Opioid-Related/Overdose-Information-Network-Data-CY-January-2018-/hbkk-dwy3/about_data). While ODIN data are updated continually, the public-access ODIN dataset is updated monthly, and the present study used data updated as of July 15, 2024. All ODIN data are based on law enforcement/first responder assessment and report.

ODIN comprises overdose incidents recorded by participating law enforcement agencies who responded to the scene of a suspected overdose, as well as overdose incidents reported on a voluntary basis by other first responders (e.g., firefighters, emergency medical personnel). Due to the voluntary nature of third-party first responder reporting in the ODIN, ODIN data do not include, nor represent, all overdoses reported to emergency services in Pennsylvania. All criminal legal agencies in Pennsylvania (federal, state, and local), all Pennsylvania 911 centers, and non-criminal justice agencies and first responders with access to the Pennsylvania Justice Network portal can enter overdose incidents and naloxone administrations in ODIN (Pennsylvania State Police, n.d.). According to a 2019 ODIN report, more than 1,300 agencies across Pennsylvania had entered data in ODIN by 2019, including state, local, and federal law enforcement, sheriffs, probation, and parole officials (Rhoads, 2019). There are 1,117 law enforcement entities in Pennsylvania (JTIC, 2018), and at the time of the present study, the publicly available list of agencies recording events in ODIN included 752 law enforcement entities (Pennsylvania State Police, 2024), primarily (but not exclusively) city/township/borough police departments and state police.

### Identifying Unique Records

Considering that any overdose event (“incident”) in ODIN may have involved multiple persons who suffered an overdose at the same time and place, we used the ODIN “victim ID” code to identify unique persons in each incident. Each ODIN record in the publicly available dataset (representing one row) can accommodate only *one* drug in the “suspected drug” column; in cases with more than one drug suspected in the same overdose incident/victim, each additional drug is listed in a separate record (additional row) that shares the same incident ID, victim ID, and date/time, but has a different entry in the “suspected drug” column. Therefore, for records with the same victim ID, incident ID, and date/time, we merged all data corresponding to the unique combination of victim ID and incident ID to maintain only one record for each person’s overdose incident. We coded new individual binary (yes/no) variables for each of the multiple different drugs reported for any given incident/victim ID combination to accommodate multiple drugs in a single, merged record for each incident/victim ID combination. We confirmed that all other measures examined in the study (victim ID, incident ID, date/time, age, gender, race/ethnicity, naloxone administration status, and survival status) were identical for our merged records.

We did not locate any instances of the same victim ID with different incident IDs (which would represent the same person experiencing separate overdose incidents recorded in the ODIN). Considering that repeat overdoses are relatively common, the lack of multiple incident IDs for the same victim ID suggested that some of the same individuals may have been assigned different victim IDs on different occasions in the public-access ODIN; as such, we consider our analyses to be at the overdose *event* level, rather than the *individual* level.

### Analytic Sample and Measures

The data used in the present study (Pennsylvania State Police, 2024) spanned January 1, 2018-July 15, 2024 and included 63 (of 67) Pennsylvania counties. Although data from each of Pennsylvania’s 67 counties have been recorded in ODIN, the present study excluded records from four counties due to limited data. We excluded Philadelphia County (which accounted for 1,242 [5%] of the ODIN opioid overdoses) because the only listed ODIN-reporting agencies from Philadelphia County were Drexel University Police and Philco; without the Philadelphia Police Department as an ODIN reporter, we considered the limited ODIN data from Philadelphia not comparable with the data from the other counties whose county/city/borough/township police forces were ODIN reporters. We also excluded three counties (Cameron, Forest, and Sullivan) with populations below 7,000 and relatively low numbers (0, 6, and 3, respectively) of ODIN-recorded opioid overdoses.

The study’s analytic sample comprised 26,635 suspected opioid-involved overdoses (each with a unique victim ID/incident ID combination). We identified opioid-involved overdoses via any opioid-related ODIN category (heroin, fentanyl, carfentanil, fentanyl analog/other synthetic opioid, pharmaceutical opioid, methadone, or suboxone) recorded as a “suspected drug.” *Suspected drugs* in each ODIN overdose report are determined by law enforcement/first responders based on sources such as interviews with the person who overdosed or others present, drug field testing, and other evidence (e.g., drugs, paraphernalia, identifiable markings from stamp bags; JTIC, 2018) at the scene of the overdose (Barboza & Angulski, 2020).

Our primary *outcome* of interest was whether the overdose was non-fatal or fatal (binary indicator for “survived”), as reported in the ODIN system based on information available to the first responder at the scene of overdose. We also examined a binary indicator for whether naloxone was administered to the individual at the scene of overdose by a first responder or bystander, as reported in the ODIN system. Our two primary *exposures* of interest comprised binary indicators for whether A) cocaine or B) methamphetamine, respectively, were listed as one of the drugs “suspected” to be co-involved in the opioid overdose (based on law enforcement/first responder assessment and report). We also examined available demographic measures, which were also based on law enforcement/first responder assessment/report (Holmes et al., 2022): the individual’s age (categories available in ODIN: 0-9, 10-19, 20-29, 30-39, 40-49, 50-59, 60-69, and 70+), gender (man, woman, “unknown”), race (White, Black, Hispanic, American Indian, Asian, or unknown) and ethnicity (Hispanic, Non-Hispanic, or unknown). In addition, we examined the county where the overdose occurred and the year of occurrence, as well as other “suspected drug” categories (binary indicators) recorded: heroin, fentanyl, carfentanil, other synthetic opioids/fentanyl analogs, pharmaceutical opioids, benzodiazepines, and alcohol.

### Statistical Analyses

We first examined suspected opioid-stimulant co-involvement in ODIN-recorded overdoses over time and across counties. We used line graphs to depict percentages of opioid overdoses that also reportedly co-involved cocaine or methamphetamine, respectively, per year, and used choropleth maps to depict these percentages at the county level. We also used descriptive statistics to summarize (and Pearson’s chi-square tests to compare) the characteristics of suspected opioid overdoses with and without cocaine and methamphetamine also co-involved, separately for a) opioid overdoses with versus without cocaine and b) opioid overdoses with versus without methamphetamine.

Next, we focused on naloxone administration and survival in suspected opioid overdoses with and without cocaine and methamphetamine co-involved. We used flow charts to depict: the proportion of opioid overdoses with and without suspected involvement of stimulants (cocaine or methamphetamine, respectively); the proportions of each of these types of overdoses in which naloxone was or was not administered; and the proportions of survival and death in each of these types of overdoses with, versus without, naloxone administered.

Finally, we conducted mediation analyses, considering overdose survival as our outcome, naloxone administration as mediator, and suspected cocaine or methamphetamine co-involvement (respectively) as the exposure variable. Mediation analyses facilitate a formal examination of potential pathways linking variables with temporal precedence (one following the other in time), allowing us to estimate what proportion of the association between stimulant co-involvement and overdose survival might be attributed to naloxone administration. Naloxone’s conceptualization as a mediator was supported by its status as an opioid overdose reversal medication used to restore breathing and prevent death after an opioid overdose. We used Stata/MP 18.5’s *mediate* suite of commands (StataCorp, 2023) based on the potential-outcomes framework (Imai et al., 2011; MacKinnon et al., 2020). This mediation approach is comparable to Baron and Kenny’s (1986) classical mediation in the context of a linear model with no exposure-mediator interaction, yet the *mediate* command provides additional flexibility and allowed us to examine a binary mediator and binary outcome (with a logit specification) and model an exposure-mediator (stimulant involvement × naloxone administration) interaction (based on prior research; Shastry et al., 2024). We examined two separate mediation models, one with cocaine co-involvement as exposure, and the other with methamphetamine co-involvement as exposure, with both models including controls for age, gender, race, county, year (categorical), and other drug co-involvement (with each drug as a binary indicator). Listwise deletion was used for the 6.2% of records with missing data on any measure; no data were missing on naloxone administration, and 4.4% of data were missing on survival status. We expressed results in terms of “total effect,” “average direct effect,” “average indirect effect,” and proportion mediated (the indirect effect as a percentage of the total effect). Although we use terms such as “direct effect” and “indirect effect,” as conventional in mediation analyses, our results are not causal in nature.

### Sensitivity Analyses

Stimulant co-use may be more common in individuals who use fentanyl, as opposed to other opioids (Daniulaityte et al., 2020), and fentanyl presents heightened overdose risks. Therefore, we conducted additional analyses focusing on suspected fentanyl-related overdoses (involving fentanyl, carfentanil, or other fentanyl analogs/synthetic opioids) instead of opioid overdoses in general. In these supplemental analyses, we compared fentanyl-stimulant and fentanyl-no-stimulant overdoses in terms of naloxone administration and survival; we also repeated the mediation models with this subset of overdoses involving fentanyl/other synthetic opioids. We also conducted supplemental analyses that repeated the mediation models without eliminating the records from Philadelphia County. Finally, we repeated our mediation models with two additional specifications in order to assess the robustness of results in two relatively extreme scenarios (Carpenter & Smuk, 2023): a) with all cases missing a response on overdose survival (4.4% of cases) imputed as “survived;” and b) with all cases missing a response on overdose survival (4.4% of cases) imputed as “died.”

## Results

### Time and County Differences in Suspected Stimulant Co-Involvement in Opioid Overdoses

As depicted in Figure 1, the proportion of ODIN-recorded opioid overdoses that reportedly co-involved cocaine rose from 3.3% in 2018 to 10.4% in the first half of 2024, and the proportion of the opioid overdoses reportedly co-involving methamphetamine increased from 1.8% in 2018 to 7.0% in the first half of 2024. The percentage of ODIN-recorded opioid overdoses co-involving cocaine or methamphetamine, as shown in Figure 2, varied widely between counties, ranging from 0.0% to 12.5% co-involving cocaine and 0.0% to 24.2% co-involving methamphetamine. Cumberland, Dauphin, and Franklin counties (located in the south-central region of the state) had the highest percentages of suspected cocaine co-involvement, while Susquehanna, Venango, and Warren counties (located across the north of the state) had the highest percentages of suspected methamphetamine co-involvement.

**Figure 1.**
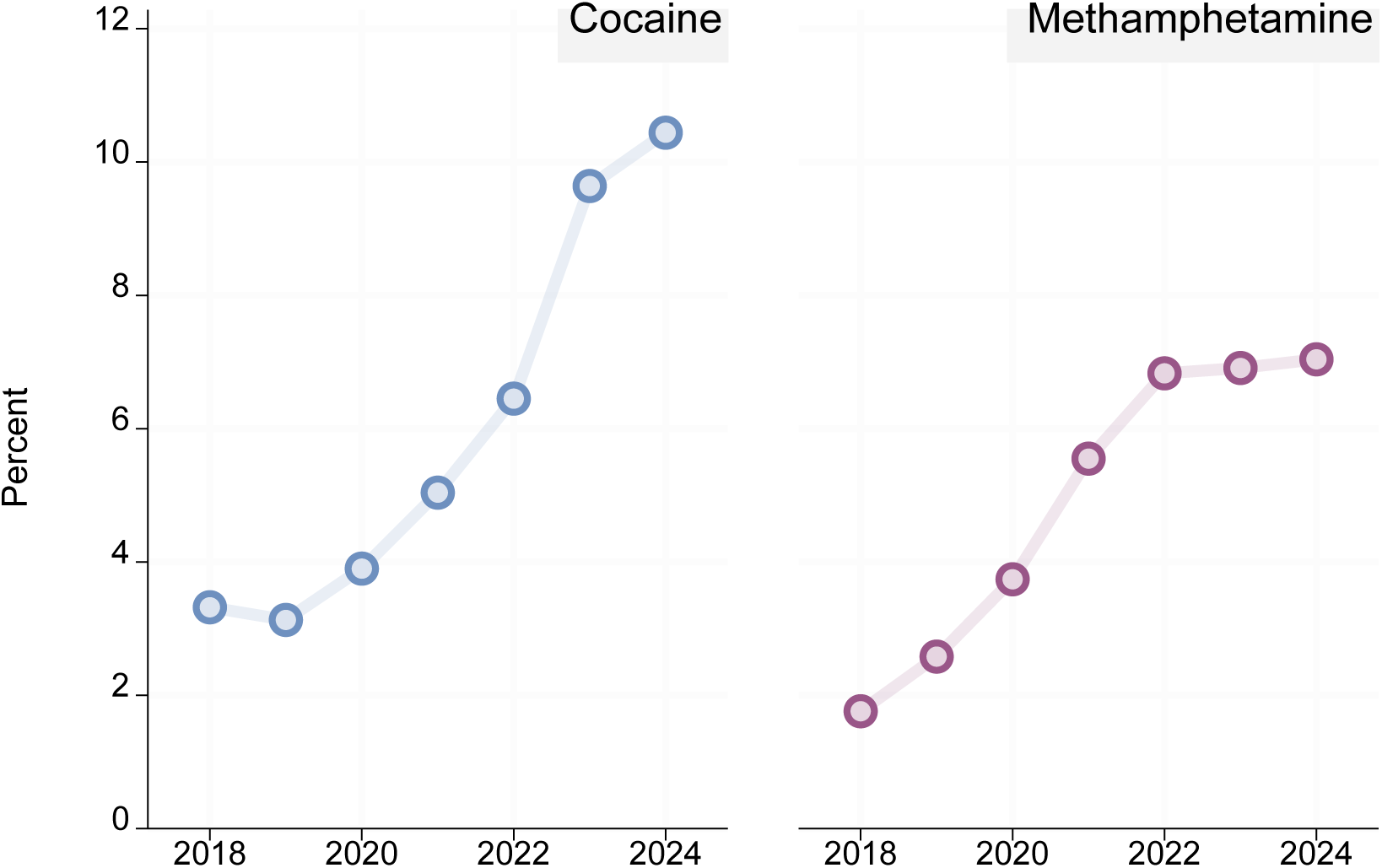
Percent of suspected opioid-related overdoses also reportedly involving (a) cocaine or (b) methamphetamine, in Pennsylvania’s Overdose Information Network (ODIN), 2018-2024.^1^. *Note.* ^1^Data do not include Philadelphia, Cameron, Forest, and Sullivan counties. Data for 2024 are through July 15, 2024.

**Figure 2.**
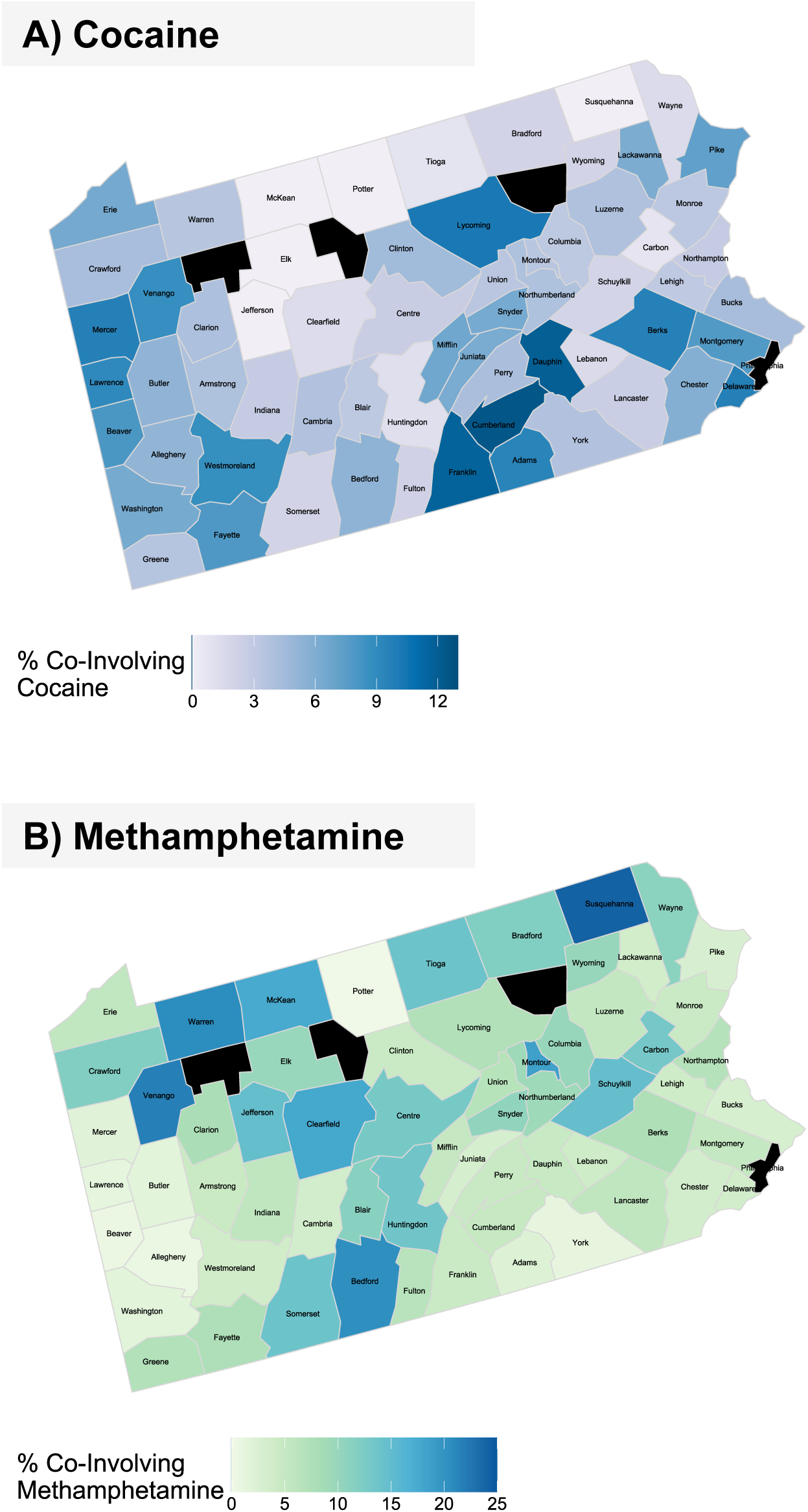
Percentages of ODIN-recorded suspected opioid overdoses reportedly co-involving cocaine or methamphetamine, 2018-2024, by county. *Notes*. Data are from the Pennsylvania Overdose Information Network (ODIN) from January 1, 2018-July 15, 2024. Counties in black (Philadelphia, Cameron, Forest, and Sullivan) are not included in the analyses.

### ODIN Overdoses Involving Opioids with or without Stimulants

#### Demographics

Table 1 summarizes characteristics of ODIN-recorded overdoses reportedly involving opioids with vs. without cocaine, as well as overdoses involving opioids with vs. without methamphetamine. Approximately two thirds of the overdoses occurred in men, ranging from 65.4% men in the opioid-cocaine group to 69.9% men in the opioid-no-cocaine group. Age distributions differed by suspected stimulant co-involvement, although overdoses were concentrated in ages 30-39 in all groups examined. White individuals represented 94.7% of the suspected opioid-methamphetamine overdoses, compared to 88.6% of the opioid-no-methamphetamine overdoses. Black individuals accounted for 17.4% of the suspected opioid-cocaine overdoses, compared to 4.3% of the opioid-methamphetamine overdoses.

**Table 1.** Selected characteristics of suspected opioid-related overdoses (*n*=26,635) reportedly co-involving cocaine or methamphetamine in Pennsylvania’s Overdose Information Network (ODIN),^1^ 2018-2024.

| | Opioid & cocaine<br>($n=1,490$) | Opioid, no cocaine<br>($n=25,145$) | P value | Opioid & metham-<br>phetamine<br>($n=1,236$) | Opioid, no methamphet-<br>amine<br>($n=25,399$) | P value |
| --- | --- | --- | --- | --- | --- | --- |
| **Age, years** |  |  | <0.001 |  |  | <0.001 |
| 0-9 | 0.2 | 0.2 |  | 0.3 | 0.2 |  |
| 10-19 | 0.9 | 1.4 |  | 1.1 | 1.4 |  |
| 20-29 | 20.5 | 27.7 |  | 22.3 | 27.5 |  |
| 30-39 | 34.2 | 38.3 |  | 39.6 | 38.0 |  |
| 40-49 | 22.6 | 18.5 |  | 21.6 | 18.6 |  |
| 50-59 | 15.0 | 9.7 |  | 11.7 | 9.9 |  |
| 60-69 | 6.2 | 3.7 |  | 3.4 | 3.9 |  |
| 70+ | 0.5 | 0.5 |  | 0.2 | 0.6 |  |
| **Gender** |  |  | <0.001 |  |  | 0.991 |
| Man | 65.4 | 69.9 |  | 69.8 | 69.7 |  |
| Woman | 34.6 | 29.8 |  | 29.9 | 30.1 |  |
| Unknown | 0.1 | 0.3 |  | 0.2 | 0.3 |  |
| **Race** |  |  | <0.001 |  |  | <0.001 |
| White | 80.5 | 89.4 |  | 94.7 | 88.6 |  |
| Black | 17.4 | 8.4 |  | 4.3 | 9.2 |  |
| AI | 0.1 | 0.1 |  | 0.2 | 0.1 |  |
| Asian | 0.7 | 0.3 |  | 0.1 | 0.3 |  |
| Unknown | 1.3 | 1.8 |  | 0.7 | 1.8 |  |
| **Ethnicity** |  |  | <0.001 |  |  | <0.001 |
| Hispanic | 7.1 | 5.6 |  | 3.4 | 5.8 |  |
| Non-Hispanic | 87.5 | 86.5 |  | 94.6 | 86.2 |  |
| Unknown | 5.4 | 7.8 |  | 2.0 | 8.0 |  |
| **Year of incident** |  |  | <0.001 |  |  | <0.001 |
| 2018 | 10.1 | 17.5 |  | 6.5 | 17.6 |  |
| 2019 | 8.5 | 15.7 |  | 8.5 | 15.6 |  |
| 2020 | 11.7 | 17.2 |  | 13.6 | 17.0 |  |
| 2021 | 12.8 | 14.2 |  | 16.9 | 14.0 |  |
| 2022 | 14.5 | 12.5 |  | 18.5 | 12.3 |  |
| 2023 | 30.8 | 17.1 |  | 26.6 | 17.5 |  |
| 2024<sup>a</sup> | 11.5 | 5.9 |  | 9.4 | 6.0 |  |
| **Other drugs co-involved** |  |  |  |  |  |  |
| Heroin | 55.6 | 80.3 | <0.001 | 57.5 | 80.0 | <0.001 |
| Fentanyl | 63.9 | 38.5 | <0.001 | 60.5 | 38.9 | <0.001 |
| Carfentanil | 1.1 | 1.0 | 0.623 | 0.7 | 1.0 | 0.404 |
| Other synth. opioids | 10.5 | 6.5 | <0.001 | 13.8 | 6.3 | <0.001 |
| RX opioids | 4.8 | 6.8 | 0.002 | 4.9 | 6.8 | 0.008 |
| Benzodiazepines | 4.6 | 1.9 | <0.001 | 4.2 | 2.0 | <0.001 |
| Alcohol | 8.4 | 2.2 | <0.001 | 3.6 | 2.5 | 0.016 |
| **Naloxone administered?** |  |  | <0.001 |  |  | <0.001 |
| Yes | 55.1 | 72.2 |  | 52.4 | 72.1 |  |
| No | 44.9 | 27.9 |  | 47.6 | 27.9 |  |
| **Survived overdose?** |  |  | <0.001 |  |  | <0.001 |
| Yes | 54.6 | 77.6 |  | 52.4 | 77.5 |  |
| No | 41.3 | 18.0 |  | 42.9 | 18.1 |  |
| Unknown | 4.1 | 4.4 |  | 4.8 | 4.4 |  |

Notes. <sup>1</sup>Data do not include Cameron, Forest, Philadelphia, and Sullivan counties. <sup>a</sup>Denotes partial year. P values based on Pearson's chi-squared tests. *Abbreviations.* AI, American Indian; Synth., synthetic; RX, pharmaceutical.

#### Other drug co-involvement

Both heroin and prescription opioids were reported in higher proportions of the overdoses without cocaine or methamphetamine reportedly involved, relative to the opioid-cocaine and opioid-methamphetamine overdoses. Conversely, fentanyl, other synthetic opioids, benzodiazepines, and alcohol were recorded in higher proportions of the suspected opioid-cocaine and opioid-methamphetamine groups compared to the opioid-no-cocaine and opioid-no-methamphetamine groups.

#### Naloxone administration and survival rates

Naloxone was reportedly administered in 72.2% of the suspected opioid-no-cocaine cases, compared to 55.1% of the opioid-cocaine cases; similarly, naloxone was administered in 72.1% of the opioid-no-methamphetamine cases, versus 52.4% of the opioid-methamphetamine cases. Finally, 18.0% of the opioid-no-cocaine overdoses ended in death, compared to 41.3% of the opioid-cocaine overdoses; 18.1% of the opioid-no-methamphetamine overdoses ended in death, versus 42.9% of the opioid-methamphetamine overdoses.

Figure 3 depicts the percentages of ODIN overdoses that reportedly ended in survival, death, or an unknown outcome, in cases where naloxone was or was not administered at the scene, separated by suspected/reported opioid-stimulant co-involvement category (opioid-cocaine vs. opioid-no-cocaine, and opioid-methamphetamine vs. opioid-no-methamphetamine). In cases with naloxone administered, more than three of every four overdoses resulted in survival, with percentages ranging from 76.2% in the suspected opioid-methamphetamine group and 80.3% in the opioid-cocaine group to 88.5% and 88.4% survival in the opioid-no-methamphetamine and opioid-no-cocaine groups, respectively. When naloxone was not administered, survival percentages differed by stimulant involvement; for example, death was the recorded outcome in 74.0% of the opioid-cocaine overdoses without naloxone administered, compared to 47.3% of the opioid-no-cocaine overdoses without naloxone administered.

**Figure 3.**
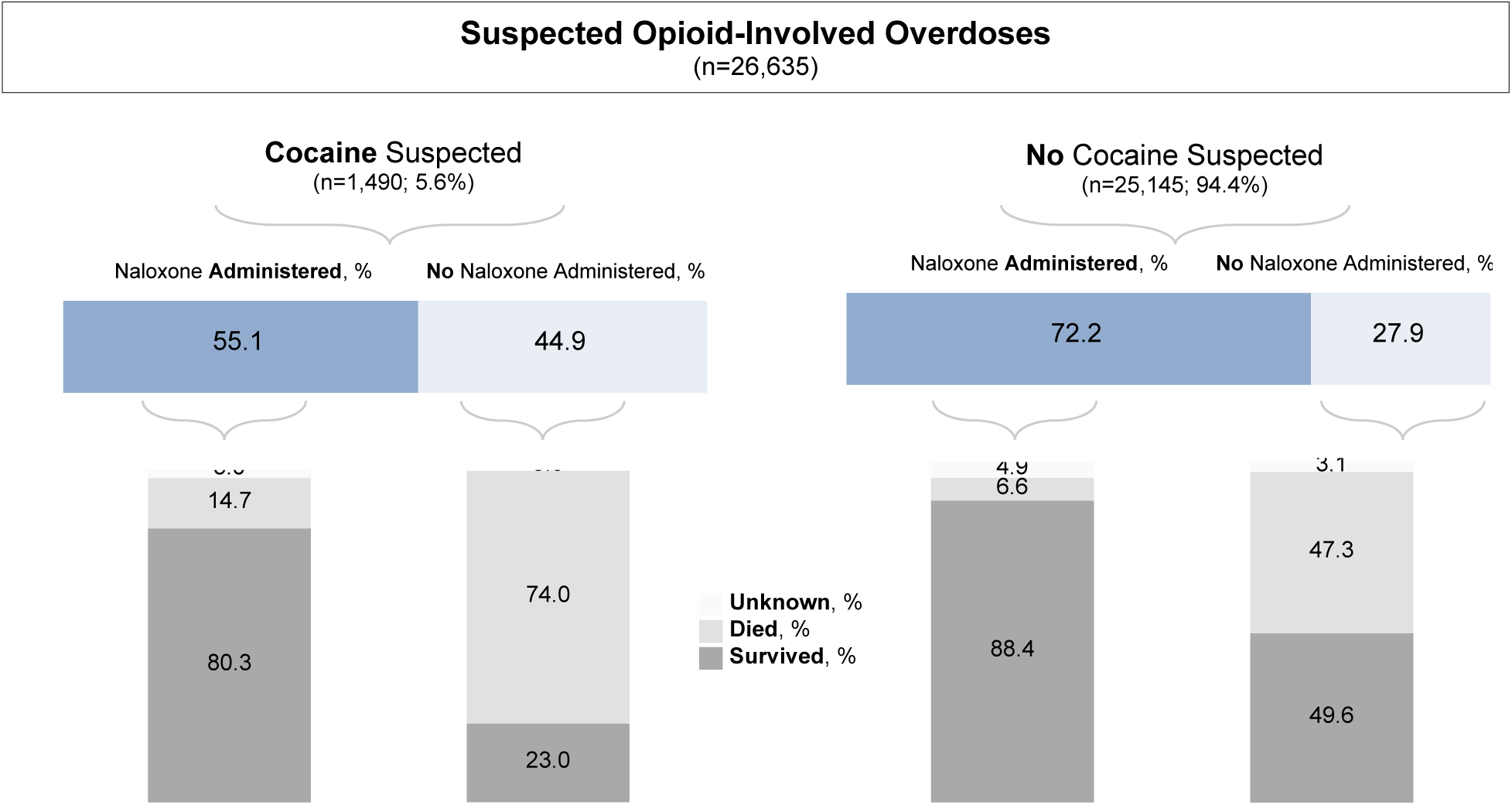
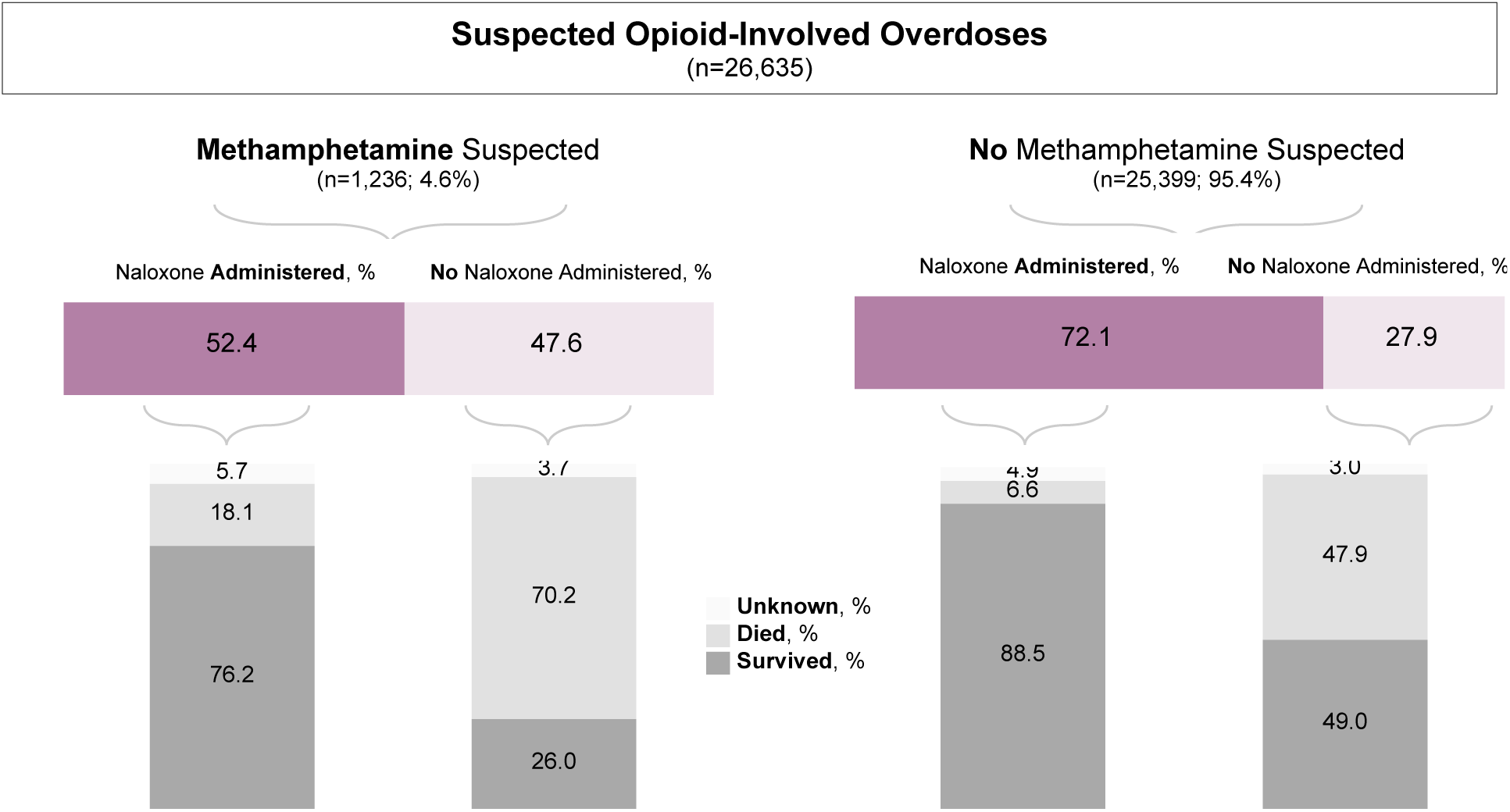
Percentages of ODIN overdoses that ended in survival, death, or an unknown outcome, by naloxone administration and reported opioid-stimulant co-involvement category. *Note*. Data do not include Philadelphia, Forest, Cameron, and Sullivan counties. Totals may not amount to 100 due to rounding.

### Mediation Analyses

Table 2 provides results from the mediation models examining naloxone administration as a mediator in the association between suspected a) cocaine or b) methamphetamine co-involvement and survival, adjusting for age, race, gender, year, county, and other drug co-involvement, also accounting for the exposure-mediator interaction. Results were similar in the a) cocaine and b) methamphetamine models: naloxone administration mediated 38.7% (95% CI, 31.3%-46.0%) of the “effect” of suspected cocaine co-involvement on survival, and 39.2% (95% CI, 31.3%-47.1%) of the “effect” of methamphetamine co-involvement on survival.

**Table 2.** Results from mediation analyses modeling naloxone administration as a mediator in the relationship between suspected (A) cocaine or (B) methamphetamine co-involvement and overdose survival, for 24,992 ODIN-recorded opioid overdoses from 63 Pennsylvania counties, 2018-2024.

|  | (A) Cocaine | (B) Methamphetamine |
| --- | --- | --- |
|  | Logit Coefficient [95% CI] | Logit Coefficient [95% CI] |
| Total effect | -0.161*** [-0.185, -0.136] | -0.166*** [-0.192, -0.139] |
| Average direct effect | -0.099*** [-0.118, -0.079] | -0.101*** [-0.124, -0.078] |
| Average indirect effect | -0.062*** [-0.077, -0.048] | -0.065*** [-0.080, -0.050] |
|  | Proportion [95% CI] | Proportion [95% CI] |
| Proportion mediated via naloxone administration | **0.387\*\*\* [0.313, 0.460]** | **0.392\*\*\* [0.313, 0.471]** |

Note. Proportion mediated denotes the indirect effect as a percentage of the total effect. Outcome equation includes exposure-mediator interaction. Covariates in outcome and mediator equations: year, incident county, age, gender, race, and drugs co-involved (fentanyl, carfentanil, other synthetic opioids/fentanyl analogs, alcohol, benzodiazepines, heroin, and methamphetamine [in cocaine model]/cocaine[in methamphetamine model]). For convention, the term "effect" is used; nevertheless, results are not causal in nature. \*\*\* $p < 0.001$ ; $p$ values based on robust standard errors. Data do not include Cameron, Forest, Philadelphia, and Sullivan counties.

### Results of Sensitivity/Supplemental Analyses

Supplemental Tables S1-S4 detail the analyses focusing on suspected *fentanyl-related* overdoses (instead of opioid overdoses broadly). Naloxone was reportedly administered in 49.9% of the suspected fentanyl-cocaine overdoses and 69.0% of the fentanyl-no-cocaine overdoses. Similarly, naloxone was reportedly administered in 48.3% of the fentanyl-methamphetamine overdoses and 68.8% of the fentanyl-no-methamphetamine overdoses. In these analyses of fentanyl-related overdoses, naloxone administration mediated 46.3% (95% CI, 36.6%-56.0%) of the relationship between cocaine co-involvement and survival, and 42.5% (95% CI, 33.1%-51.9%) of the relationship between methamphetamine co-involvement and survival. As presented in Supplemental Tables S5-S7, mediation results were nearly identical to main results (Table 2) even when including cases from Philadelphia County (Table S5) or imputing all missing survival responses (4.4% of cases) as either “survived” (Table S6) or “died” (Table S7) instead of eliminating the missing responses (as in Table 2).

## Discussion

Using data from 26,635 law enforcement-attended opioid overdose events recorded in Pennsylvania’s Overdose Information Network (ODIN), the present study explored the relationship between naloxone administration and survival in cases of overdose involving opioids with or without stimulants co-involved. Study results indicated that rates of naloxone administration and survival were notably lower in suspected opioid-stimulant overdoses than in opioid-no-stimulant overdoses, even in analyses adjusting for the co-involvement of other drugs such as fentanyl and fentanyl analogs, benzodiazepines, and alcohol.

Although stimulants are co-involved in only a portion of opioid-related overdoses, this proportion is increasing over time (Hoots et al., 2020; Spencer et al., 2023). In our study, the proportion of ODIN-recorded opioid overdoses with cocaine reportedly co-involved rose from 3.3% in 2018 to 10.4% in the first half of 2024, and the proportion of opioid overdoses with methamphetamine reportedly co-involved increased from 1.8% in 2018 to 7.0% in the first half of 2024. In consideration of increases in opioid-stimulant co-use nationwide (Jones et al., 2020), our study’s finding of higher probabilities of death (and lower probabilities of naloxone receipt) among opioid-stimulant overdoses emphasizes the importance of prioritizing and tailoring overdose prevention and response efforts for opioid-stimulant co-exposure.

### Lower Rates of Naloxone Administered in ODIN Opioid-Stimulant-Involved Overdoses

In the ODIN-recorded overdoses in the present study, naloxone was reportedly administered in 72.2% of the overdoses that involved opioids without cocaine, compared to 55.1% of the overdoses involving opioids with cocaine; similarly, naloxone was administered in 72.1% of the overdoses involving opioids without methamphetamine, versus 52.4% of the overdoses involving opioids with methamphetamine. Although our study was unable to examine reasons for this finding, prior research suggests several plausible explanations. Since the clinical presentation and symptoms of opioid-stimulant-involved overdoses may differ from opioid-overdoses in general (Glidden et al., 2023), in some cases of opioid-stimulant involved overdose, it may not be quickly apparent to bystanders or law enforcement whether the person is suffering from an overdose involving opioids. For example, the pupil dilation associated with some stimulants (Substance Abuse and Mental Health Services Administration, 2021) contrasts with the pupil constriction associated with opioids (Williams & Erickson, 2000). Moreover, in cases in which opioids and stimulants may have been used at different times throughout the day, or in cases of fentanyl-contaminated stimulants (Daniulaityte et al., 2023; Wagner et al., 2023), evidence of opioids (which may prompt bystanders to administer naloxone) may not be readily visible. Although all overdose events examined in the present study reportedly involved opioids, it is not clear what proportion evidenced symptoms of opioid-induced respiratory depression for which naloxone is indicated.

Individuals who primarily use stimulants may be less likely to carry take-home naloxone (Hughto et al., 2022) that could be administered by a bystander even before the arrival of first responders. It is also possible that some responding law enforcement officers may have heightened concerns in stimulant-involved cases that may reduce their likelihood of prioritizing administering naloxone (e.g., beliefs about aggressive behavior related to methamphetamine, doubts about naloxone or their role in administering it; Bucerius et al., 2022; Murphy & Russell, 2020; Silverstein et al., 2024; Wootson, 2017). When responding to an overdose scene, some local law enforcement officers may recognize an individual from a prior contact, and the officer’s previous experiences or perceptions about the person’s substance use practices (e.g., whether the person typically uses stimulants rather than opioids, whether the person has experienced a prior opioid overdose) may influence in-the-moment decisions about administering naloxone. Finally, in some ODIN overdose cases in which naloxone was not administered, the overdose victim may have already been deceased by the time a first responder arrived on scene.

### Lower Rates of Survival in ODIN Opioid-Stimulant-Involved Overdoses

Prior studies have documented higher rates of overdose mortality in persons with concurrent stimulant and opioid use disorders, relative to one disorder only (Colell et al., 2018; Krawczyk et al., 2020), and a recent cohort study from Canada found twice the hazard of fatal overdose among individuals with opioid and stimulant use, compared to opioid use alone (Palis et al., 2022). Consistent with these results, in the ODIN-recorded overdoses examined in the present study, higher proportions of opioid-stimulant overdoses were fatal (i.e., 42.9% of the opioid-methamphetamine overdoses ended in death, compared to 18.1% of the opioid-no-methamphetamine overdoses, and 41.3% of the opioid-cocaine overdoses ended in death, compared to 18.0% of the opioid-no-cocaine overdoses). In mediation models, naloxone administration mediated approximately 39% of the association between stimulant co-involvement and survival, after adjusting for age, race, gender, year, county, and other drug co-involvement. Notably, these patterns were generally consistent even when examining stimulant co-involvement in fentanyl-related overdoses specifically rather than opioid-related overdoses in general.

Beyond differences in naloxone administration, numerous other factors may potentially contribute to lower survival in opioid-stimulant overdoses. While naloxone addresses opioid-induced respiratory depression, no comparable reversal agent is available to specifically counteract stimulant effects, and stimulant health impacts are often chronic and cumulative (Riley et al., 2022). It is also possible that some individuals who co-use opioids and stimulants may use higher opioid quantities as stimulant effects may temporarily mask some opioid effects (Shastry et al., 2024). Qualitative research indicates that some individuals who use drugs believe that methamphetamine can be used to prevent or reverse an opioid overdose (Daniulaityte et al., 2022). As such, it is also possible that some opioid-methamphetamine ODIN overdoses represented instances in which: a) an individual believed that use of methamphetamine would reduce their opioid overdose risk; or b) a peer injected methamphetamine in someone experiencing fentanyl toxicity as part of an attempt to reverse the overdose (Daniulaityte et al., 2022).

Although *intentional co-use* of opioids and stimulants is likely more common than exposure to stimulants that have been *contaminated* by fentanyl (Jones et al., 2020), fentanyl was detected in 12-15% of powder cocaine or methamphetamine samples analyzed in a cross-state US drug checking program from 2021-2023 (Wagner et al., 2023). Risk of fatality from fentanyl-contaminated stimulants may be particularly pronounced in individuals who primarily use stimulants, since naloxone carriage rates and opioid tolerance are lower in these individuals compared to persons who regularly use opioids (Hughto et al., 2022).

The overdose survival differences observed in the present study may also be influenced by socioeconomic, racial, and geographic disparities associated with stimulant use. For instance, methamphetamine-involved overdose deaths are disproportionately observed in rural areas of the US (Hedegaard & Spencer, 2021), and a recent nationwide analysis found that counties with the highest psychostimulant (e.g., methamphetamine) and opioid mortality rates generally had higher Social Vulnerability Index scores (Segel et al., 2024). Furthermore, cocaine-involved overdose deaths disproportionately impact Black Americans (Cano et al., 2020, 2022; Jones et al., 2023a), and exposure to fentanyl-contaminated stimulants has been identified as a risk disproportionately impacting Black populations (Ray et al., 2020; Shufflebarger et al., 2024). In the present study, Black individuals accounted for approximately one in six of the opioid-cocaine involved ODIN overdoses, compared to one in twelve of the opioid-no-cocaine overdoses. In addition to disparities in treatment (Barnett et al., 2023; Shufflebarger et al., 2024), Black Americans experience pronounced disparities in drug arrests (Mitchell & Caudy, 2015) which can in turn contribute to hesitancy toward contacting 911 in case of overdose (Watson et al., 2018) as well as increased risk of overdose post-release (Joudrey et al., 2019). National research also indicates that overdose deaths in the Black population are overrepresented among relatively older (e.g., 55-64) age groups (Jones et al., 2023b); age (as well as health status) is considered a particularly salient risk factor in stimulant-involved deaths (Riley et al., 2022).

Prior studies have indicated that individuals who co-use opioids and stimulants are more likely to suffer from poor health and comorbidities (relative to those who use opioids only; Palis et al., 2022), and chronic use of stimulants can contribute to the development of conditions such as hypertension, atherosclerosis, and cardiovascular disease that may leave individuals more vulnerable to overdose death (Riley et al., 2022). Finally, persons who co-use opioids and stimulants may also face additional stigma and reduced engagement with healthcare systems, as well as lower receipt of medications for opioid use disorder that reduce the risk of fatal overdose (Palis et al., 2022). Although not directly tested in the present study, it is plausible that the disparities and marginalization associated with some types of stimulant or opioid-stimulant use (Daniulaityte et al., 2020; Jones et al., 2023c) may contribute to the elevated fatality rates observed in the opioid-stimulant ODIN overdoses.

## Limitations

The data used in this study include only overdose cases in which an ODIN-participating law enforcement agency or other first responder was present at the scene. As such, the data do not include overdose cases in which law enforcement did not respond (e.g., overdoses which were reversed without calling 911) or cases with only first responders who were not ODIN-reporting agencies. The extent to which different agencies report events in ODIN varies between counties and over time, and although we were able to adjust for county and year in analyses, we were not able to adjust for sub-county area or type of ODIN reporting agency (e.g., law enforcement versus non law enforcement) due to lack of these measures. Since the Philadelphia Police Department was not listed as an ODIN-reporting agency at the time of the study, all analyses excluded Philadelphia County, the county with the second-highest overdose death rate in the state (according to 2022 data; CDC, 2024a). Philadelphia County is the largest county in the state and a particularly racially and ethnically diverse area (Pennsylvania State Data Center, 2022), but historically, Philadelphia County’s overdose data has been subject to notable limitations; for example, according to a CDC publication, in 2017, 95% of drug overdose death certificates from Philadelphia County were missing the specific name of the drug involved in the death, compared to less than 2% missing this information in Allegheny County (Jones et al., 2019), Pennsylvania’s second-largest county.

All measures in the present study (including demographics, suspected drugs involved, and overdose fatality or survival) were based on the report of law enforcement or other first responders, using information available at the overdose scene and at the time of the overdose (rather than information from future follow-up). Because they are based on first responder report, ODIN demographics such as race/ethnicity may not always match the individual’s self-identified demographic group; at the same time, these measures potentially reflect how first responders *perceive* an individual, and this perception has the potential to influence how the individual may be treated.

In the absence of data from formal toxicology, ODIN drug involvement measures only reflect the drugs *suspected* to be involved in the overdose based on first responder investigation. It is plausible that *fatal* overdoses may have received more investigative time and resources from law enforcement than overdoses that were *non-fatal*, which could potentially render the information recorded on fatal overdoses (such as the drugs suspected to be involved) more complete. Many measures relevant to overdose survival were not available in the dataset; as such, we were unable to account for factors such as the individual’s health status, drug use history, quantity of drug used, drug purity and composition, route of drug administration, use of drugs with others or alone, emergency response time, and other emergency care provided (e.g., CPR or rescue breathing).

We modeled naloxone administration as a potential mediator in the relationship between suspected stimulant co-involvement and survival, based on the following conceptualized steps: 1) a person has an overdose involving either opioids and stimulants or opioids without stimulants; 2) a bystander or first responder subsequently administers, or does not administer, naloxone; and 3) the person either subsequently recovers or dies. However, ODIN does not indicate timing of death, so we were unable to distinguish between deaths that occurred before first responders (or even other bystanders) arrived, versus deaths that occurred while first responders were on the scene. We have no evidence to suggest that ODIN opioid-stimulant and opioid-no-stimulant cases systematically differed in timing of death and first responder arrival, but if systematic differences existed, this could bias analyses of naloxone administration and survival. For example, hypothetically, if opioid-stimulant cases were more likely than opioid-no-stimulant cases to end in death before a first responder’s or bystander’s arrival, then this could contribute to lower naloxone administration rates in the opioid-stimulant group if naloxone was not administered because the person had already passed away. Moreover, if someone administered naloxone to a person who was already deceased, there would be no potential for survival even though naloxone was administered. Finally, although naloxone dosage is recorded in ODIN, we did not include dosage in our analyses in consideration of missing data on this measure, and no information was available regarding route of administration. Overall, the study results are descriptive and correlational rather than causal in nature.

## Implications and Conclusions

The present study documented notably lower rates of naloxone administration and survival in opioid-stimulant overdoses (relative to opioid-no-stimulant overdoses) in the Pennsylvania Overdose Information Network. These results underscore the importance of optimizing training and information dissemination regarding recognizing and responding to opioid-stimulant-involved overdoses. Opioid-stimulant overdose response may be strengthened through providing first responders and bystanders with training that specifically discusses opioid-stimulant-involved overdoses, as well as consistent messaging that addresses stigma and concerns regarding naloxone (Murphy & Russell, 2020). Specific information to include in first responder training on opioid-stimulant overdose response would ideally be informed by emergency medical providers as well as stakeholders such as first responders, harm reduction community response workers, and persons with lived experience, especially given concerns surrounding police involvement in overdose calls (Del Pozo, 2022; Doe-Simkins et al., 2022). At the same time, increasing harm reduction support and messaging for individuals who use drugs (informed, designed, and/or implemented by individuals with lived experience) may help bolster knowledge regarding the risks of opioid-stimulant co-exposure and strategies to minimize risk, in addition to general harm reduction recommendations such as carrying naloxone and not using drugs alone. Finally, since the study’s results highlight persons who experience opioid-stimulant overdoses as a group facing high risk of fatality, results also support upstream efforts to reduce marginalization, social inequities, and barriers to healthcare, treatment, and resources within this group.

## Data Availability

All data are publicly-available online at https://data.pa.gov/Opioid-Related/Overdose-Information-Network-Data-CY-January-2018-/hbkk-dwy3/about_data

## Funding

Funding is reported from the National Institutes of Health (PI Abenaa Jones: K01 DA051715).

## Declaration of interest

Frank LoVecchio, DO, MPH reports the following: NIH grants, speaker bureau ABBVIE for antibiotics only. No other conflicts of interest were reported.

---

**Supplemental Table S1.** Percentage of ODIN-recorded suspected opioid-involved overdoses reportedly involving fentanyl, by year.

| Year | Percent involving fentanyl |
| --- | --- |
| 2018 | 17.5 |
| 2019 | 27.1 |
| 2020 | 33.7 |
| 2021 | 42.3 |
| 2022 | 49.1 |
| 2023 | 61.2 |
| 2024 (January-July) | 64.5 |

*Notes.* Data from 63 counties (excluding Philadelphia, Forest, Cameron, and Sullivan counties). *Abbreviations:* ODIN, Overdose Information Network.

**Supplemental Table S2.** Percentages of suspected *<u>fentanyl</u>*-related overdoses (n=11,588) that reportedly ended in survival, death, or an unknown outcome, by naloxone administration and suspected *<u>cocaine</u>* co-involvement.

| Cocaine Involved? | Naloxone Administered? | % Survived | % Died | % Unknown |
| --- | --- | --- | --- | --- |
| Y | Y | 77.71 | 17.05 | 5.23 |
| Y | N | 17.76 | 80.12 | 2.12 |
| N | Y | 85.89 | 9.06 | 5.05 |
| N | N | 34.16 | 63.24 | 2.60 |

*Notes.* N=1,034 fentanyl-cocaine overdoses and 10,554 fentanyl-no-cocaine overdoses. Naloxone was reportedly administered in 49.90% of the fentanyl-cocaine overdoses and 69.02% of the fentanyl-no-cocaine overdoses. Data from Pennsylvania Overdose Information Network, 2018-July 2024. Data do not include Philadelphia, Forest, Cameron, or Sullivan counties. Totals may not amount to 100 due to rounding. Fentanyl-related refers to overdoses reportedly involving fentanyl, carfentanil, or other fentanyl analogs/synthetic opioids. *Abbreviations:* Y, yes; N, no; %, percentage

**Supplemental Table S3.** Percentages of suspected *<u>fentanyl</u>*-related overdoses (n=11,588) that reportedly ended in survival, death, or an unknown outcome, by naloxone administration and suspected *<u>methamphetamine</u>*co-involvement.

| Methamphetamine Involved? | Naloxone Administered? | % Survived | % Died | % Unknown |
| --- | --- | --- | --- | --- |
| Y | Y | 72.73 | 20.88 | 6.39 |
| Y | N | 17.47 | 78.85 | 3.68 |
| N | Y | 86.04 | 8.97 | 4.99 |
| N | N | 33.79 | 63.82 | 2.39 |

Notes. N=842 fentanyl-cocaine overdoses and 10,746 fentanyl-no-cocaine overdoses. Naloxone was reportedly administered in 48.34% of the fentanyl-cocaine overdoses and 68.80% of the fentanyl-no-cocaine overdoses. Data from Pennsylvania Overdose Information Network, 2018-July 2024. Data do not include Philadelphia, Forest, Cameron, or Sullivan counties. Totals may not amount to 100 due to rounding. Fentanyl-related refers to overdoses reportedly involving fentanyl, carfentanil, or other fentanyl analogs/synthetic opioids. Data from Pennsylvania Overdose Information Network, 2018-July 2024. Abbreviations: Y, yes; N, no; %, percentage

**Supplemental Table S4.** Results from mediation analyses modeling naloxone administration as a mediator in the relationship between suspected (A) cocaine or (B) methamphetamine co-involvement and overdose survival, for 10,975 ODIN-recorded *<u>fentanyl</u>*-related overdoses from 63 Pennsylvania counties, 2018-2024.

|  | (A) Cocaine | (B) Methamphetamine |
| --- | --- | --- |
|  | Logit Coefficient [95% CI] | Logit Coefficient [95% CI] |
| Total effect | -0.175*** [-0.206, -0.143] | -0.206*** [-0.241, -0.171] |
| Average direct effect | -0.094*** [-0.120, -0.068] | -0.119*** [-0.149, -0.088] |
| Average indirect effect | -0.081*** [-0.101, -0.061] | -0.088*** [-0.109, -0.066] |
|  | Proportion [95% CI] | Proportion [95% CI] |
| Proportion mediated via naloxone administration | **0.463\*\*\* [0.366, 0.560]** | **0.425\*\*\* [0.331, 0.519]** |

*Note.* Proportion mediated denotes the indirect effect as a percentage of the total effect. Outcome equation includes exposure–mediator interaction. Covariates in outcome and mediator equations: year, incident county, age, gender, race, and drugs suspected to be co-involved (carfentanil, other synthetic opioids/fentanyl analogs, alcohol, benzodiazepines, heroin, and methamphetamine [in cocaine model]/cocaine[in methamphetamine model]). For convention, the term “effect” is used; nevertheless, results are not causal. \*\*\* $p < 0.001$ ; p values based on robust standard errors. Data from Pennsylvania Overdose Information Network, 2018-July 2024. Data do not include Forest, Philadelphia, Cameron, and Sullivan counties. Fentanyl-related refers to overdoses reportedly involving fentanyl, carfentanil, or other fentanyl analogs/synthetic opioids.

**Supplemental Table S5.** Results from mediation analyses modeling naloxone administration as a mediator in the relationship between suspected (A) cocaine or (B) methamphetamine co-involvement and overdose survival, for 25,320 ODIN-recorded <u>opioid</u> overdoses from <u>64</u> Pennsylvania counties (<u>including</u> <u>Philadelphia County</u>), 2018-2024.

|  | (A) Cocaine | (B) Methamphetamine |
| --- | --- | --- |
|  | Logit Coefficient [95% CI] | Logit Coefficient [95% CI] |
| Total effect | -0.16 [-0.18, -0.13] | -0.16 [-0.19, -0.14] |
| Average direct effect | -0.10 [-0.12, -0.08] | -0.10 [-0.12, -0.08] |
| Average indirect effect | -0.06 [-0.08, -0.05] | -0.06 [-0.08, -0.05] |
|  | Proportion [95% CI] | Proportion [95% CI] |
| Proportion mediated via naloxone administration | **0.39 [0.31, 0.46]** | **0.39 [0.31, 0.47]** |

*Note.* Proportion mediated denotes the indirect effect as a percentage of the total effect. Outcome equation includes exposure–mediator interaction. Covariates in outcome and mediator equations: year, incident county, age, gender, race, and drugs co-involved (fentanyl, carfentanil, other synthetic opioids/fentanyl analogs, alcohol, benzodiazepines, heroin, and methamphetamine [in cocaine model]/cocaine[in methamphetamine model]). For convention, the term “effect” is used; nevertheless, results are not causal in nature. \*\*\* $p < 0.001$ ; $p$ values based on robust standard errors. Data from Pennsylvania Overdose Information Network, 2018-July 2024. Data do not include Cameron, Forest, and Sullivan counties.

**Supplemental Table S6.**
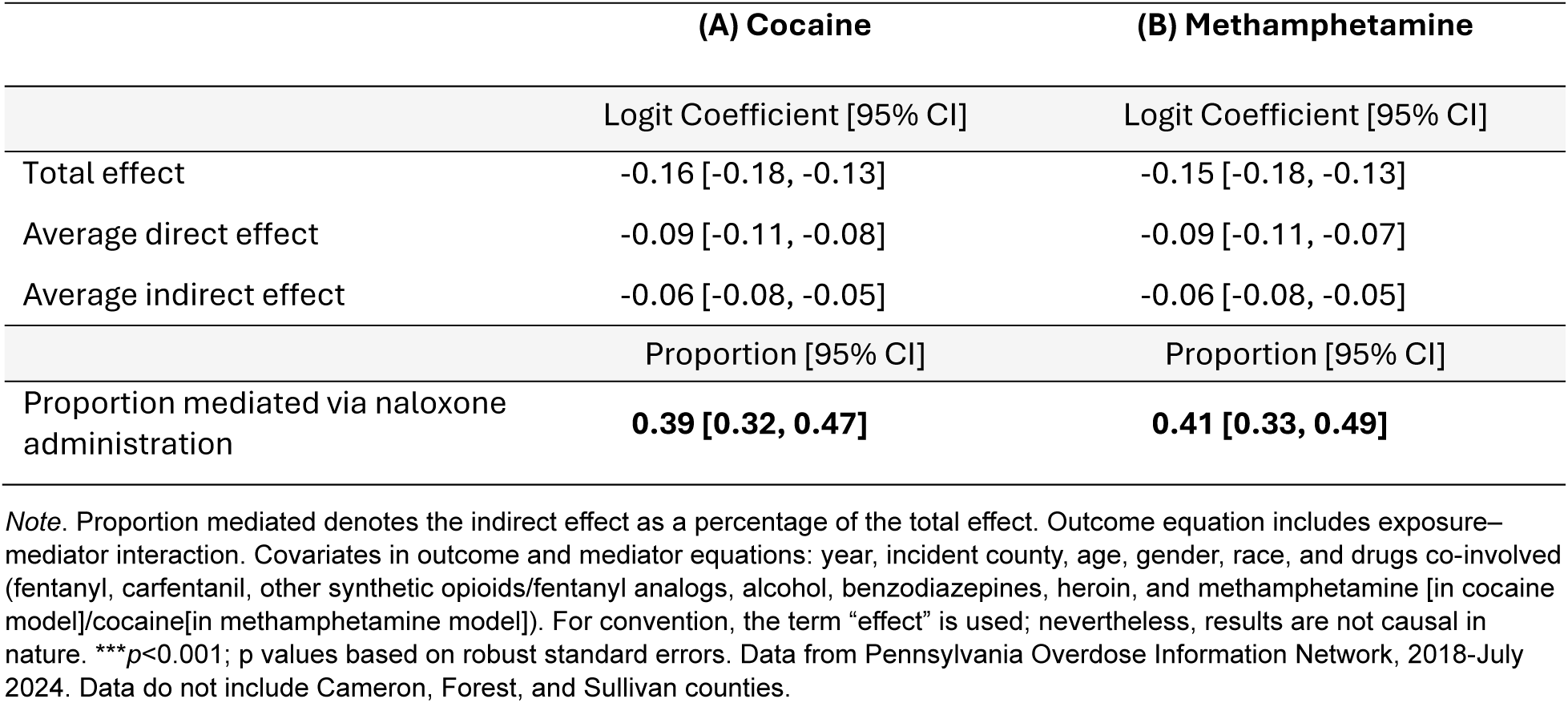
Results from mediation analyses modeling naloxone administration as a mediator in the relationship between suspected (A) cocaine or (B) methamphetamine co-involvement and overdose survival, for 26,120 ODIN-recorded opioid overdoses from 63 Pennsylvania counties, 2018-2024, <u>with all cases (4.4%) missing data on the dependent variable (overdose survival) coded as</u> <u>“survived.”</u>.

**Supplemental Table S7.** Results from mediation analyses modeling naloxone administration as a mediator in the relationship between suspected (A) cocaine or (B) methamphetamine co-involvement and overdose survival, for 26,120 ODIN-recorded opioid overdoses from 63 Pennsylvania counties, 2018-2024, <u>with all cases (4.4%) missing data on the dependent variable (overdose survival) coded as “died.”</u>.

|  | (A) Cocaine | (B) Methamphetamine |
| --- | --- | --- |
|  | Logit Coefficient [95% CI] | Logit Coefficient [95% CI] |
| Total effect | -0.16 [-0.18, -0.13] | -0.16 [-0.19, -0.14] |
| Average direct effect | -0.10 [-0.12, -0.08] | -0.10 [-0.13, -0.08] |
| Average indirect effect | -0.06 [-0.08, -0.05] | -0.06 [-0.08, -0.05] |
|  | Proportion [95% CI] | Proportion [95% CI] |
| Proportion mediated via naloxone administration | **0.39 [0.31, 0.46]** | **0.38 [0.30, 0.46]** |

*Note.* Proportion mediated denotes the indirect effect as a percentage of the total effect. Outcome equation includes exposure–mediator interaction. Covariates in outcome and mediator equations: year, incident county, age, gender, race, and drugs co-involved (fentanyl, carfentanil, other synthetic opioids/fentanyl analogs, alcohol, benzodiazepines, heroin, and methamphetamine [in cocaine model]/cocaine[in methamphetamine model]). For convention, the term “effect” is used; nevertheless, results are not causal in nature. \*\*\* $p < 0.001$ ; $p$ values based on robust standard errors. Data from Pennsylvania Overdose Information Network, 2018-July 2024. Data do not include Cameron, Forest, and Sullivan counties.

## Notes

### Author Declarations

The Institutional Review Board of Arizona State University waived ethical approval for this work, exempting the study based on Federal Regulations 45CFR46(4).

### Summary of Updates

additional background information and details regarding the data source and its use in prior literature

## References

Ahmed, S., Sarfraz, Z., & Sarfraz, A. (2022). A changing epidemic and the rise of opioid-stimulant co-use. Frontiers in Psychiatry, 13, 918197. 10.3389/fpsyt.2022.918197

Bailey, A., Andraka-Christou, B., Rouhani, S., Clark, M. H., Atkins, D., & del Pozo, B. (2024). Beliefs of US chiefs of police about substance use disorder, fentanyl exposure, overdose response, and use of discretion: Results from a national survey. Research Square. 10.21203/rs.3.rs-5333150/v1

Barboza, G. E., & Angulski, K. (2020). A descriptive study of racial and ethnic differences of drug overdoses and naloxone administration in Pennsylvania. International Journal of Drug Policy, 78, 102718. 10.1016/j.drugpo.2020.102718

Barnett, M. L., Meara, E., Lewinson, T., Hardy, B., Chyn, D., Onsando, M., Huskamp, H. A., Mehrotra, A., & Morden, N. E. (2023). Racial inequality in receipt of medications for opioid use disorder. New England Journal of Medicine, 388(19), 1779–1789. doi: 10.1056/NEJMsa2212412

Baron, R. M., & Kenny, D. A. (1986). The moderator–mediator variable distinction in social psychological research: Conceptual, strategic, and statistical considerations. Journal of Personality and Social Psychology, 51(6), 1173–1182. 10.1037/0022-3514.51.6.1173

Bucerius, S., Berardi, L., Haggerty, K. D., & Krahn, H. (2022). New drugs, new fears: Synthetic opioids and adaptations to police practice. Policing and Society, 32(7), 832–845. 10.1080/10439463.2021.2020783

Burgess-Hull, A. J., Smith, K. E., Panlilio, L. V., Schriefer, D., Preston, K. L., Alter, A., … & Epstein, D. H. (2022). Nonfatal opioid overdoses before and after Covid-19: Regional variation in rates of change. PLoS One, 17(3), e0263893. 10.1371/journal.pone.0263893

Britch, S. C., & Walsh, S. L. (2022). Treatment of opioid overdose: Current approaches and recent advances. Psychopharmacology, 239(7), 2063–2081. 10.1007/s00213-022-06125-5

Cano, M., Oh, S., Salas-Wright, C. P., & Vaughn, M. G. (2020). Cocaine use and overdose mortality in the United States: Evidence from two national data sources, 2002–2018. Drug and Alcohol Dependence, 214, 108148. 10.1016/j.drugalcdep.2020.108148

Cano, M., Salas-Wright, C. P., Oh, S., Noel, L., Hernandez, D., & Vaughn, M. G. (2022). Socioeconomic inequalities and Black/White disparities in US cocaine-involved overdose mortality risk. Social Psychiatry and Psychiatric Epidemiology, 57(10), 2023–2035. 10.1007/s00127-022-02255-5

Carpenter, J. R., & Smuk, M. (2021). Missing data: A statistical framework for practice. Biometrical Journal, 63(5), 915–947. 10.1002/bimj.202000196

Center for Rural Pennsylvania. (n.d.). Rural urban definitions. https://www.rural.pa.gov/data/rural-urban-definitions

Centers for Disease Control and Prevention. (2024a). National Center for Health Statistics Mortality Data on CDC WONDER. https://wonder.cdc.gov/mcd.html

Centers for Disease Control and Prevention. (2024b). Public Health Considerations for Strategies and Partnerships. https://www.cdc.gov/overdose-prevention/php/public-health-strategy/index.html

Ciccarone, D. (2021). The rise of illicit fentanyls, stimulants and the fourth wave of the opioid overdose crisis. Current Opinion in Psychiatry, 34(4), 344–350. doi: 10.1097/YCO.0000000000000717

Colell, E., Domingo-Salvany, A., Espelt, A., Parés-Badell, O., & Brugal, M. T. (2018). Differences in mortality in a cohort of cocaine use disorder patients with concurrent alcohol or opiates disorder. Addiction, 113(6), 1045–1055.

Davis, C. S., & Carr, D. (2020). Over the counter naloxone needed to save lives in the United States. Preventive Medicine, 130, 105932. 10.1016/j.ypmed.2019.105932

Daniulaityte, R., Ruhter, L., Juhascik, M., & Silverstein, S. (2023). Attitudes and experiences with fentanyl contamination of methamphetamine: exploring self-reports and urine toxicology among persons who use methamphetamine and other drugs. Harm Reduction Journal, 20(1), 54. 10.1186/s12954-023-00782-1

Daniulaityte, R., Silverstein, S. M., Crawford, T. N., Martins, S. S., Zule, W., Zaragoza, A. J., & Carlson, R. G. (2020). Methamphetamine use and its correlates among individuals with opioid use disorder in a Midwestern US city. Substance Use & Misuse, 55(11), 1781–1789. 10.1080/10826084.2020.1765805

Daniulaityte, R., Silverstein, S. M., Getz, K., Juhascik, M., McElhinny, M., & Dudley, S. (2022). Lay knowledge and practices of methamphetamine use to manage opioid-related overdose risks. International Journal of Drug Policy, 99, 103463. 10.1016/j.drugpo.2021.103463

Del Pozo, B. (2022). Reducing the iatrogenesis of police overdose response: Time is of the essence. American Journal of Public Health, 112(9), 1236–1238. doi: 10.2105/AJPH.2022.306987

Doe-Simkins, M., El-Sabawi, T., & Carroll, J. J. (2022). Whose concerns? It’s time to adjust the lens of research on police-involved overdose response. American Journal of Public Health, 112(9), 1239–1241.

Fredericksen, R. J., Baker, R., Sibley, A., Estadt, A. T., Colston, D., Mixson, L. S., Walters, S., Bresett, J., Levander, X. A., Leichtling, G., Davy-Mendez, T., Powell, M., Stopka, T. J., Pho, M., Feinberg, J., Ezell, J., Zule, W., Seal. D., Cooper, H. L. F., Whitney, B. M., Delaney, J. A. C., Crane, H. M., & Tsui, J. I. (2024). Motivation and context of concurrent stimulant and opioid use among persons who use drugs in the rural United States: A multi-site qualitative inquiry. Harm Reduction Journal, 21(1), 74. 10.1186/s12954-024-00986-z

Glidden, E., Suen, K., Mustaquim, D., Vivolo-Kantor, A., Brent, J., Wax, P., Aldy, K., On behalf of the Toxicology Investigators Consortium (ToxIC) Study Group. (2023). Characterization of nonfatal opioid, cocaine, methamphetamine, and polydrug exposure and clinical presentations reported to the Toxicology Investigators Consortium Core Registry, January 2010–December 2021. Journal of Medical Toxicology, 19(2), 180–189. 10.1007/s13181-022-00924-0

Hedegaard, H., & Spencer, M. R. (2021). Urban-rural differences in drug overdose death rates, 1999-2019. NCHS Data Brief, no 403. National Center for Health Statistics. 10.15620/cdc:102891

High Intensity Drug Trafficking Areas Overdose Response Strategy. (2019). 2019 annual report. https://www.hidtaprogram.org/pdf/ors_report_2019.pdf

Holmes, L. M., & King, B. H. (2023). County-level variation in synthetic opioid and heroin overdose incidents in Pennsylvania during the COVID-19 pandemic. Applied Geography, 155, 102977. 10.1016/j.apgeog.2023.102977

Holmes, L. M., Rishworth, A., & King, B. H. (2022). Disparities in opioid overdose survival and naloxone administration in Pennsylvania. Drug and Alcohol Dependence, 238, 109555. 10.1016/j.drugalcdep.2022.109555

Hoots, B., Vivolo-Kantor, A., & Seth, P. (2020). The rise in non-fatal and fatal overdoses involving stimulants with and without opioids in the United States. Addiction, 115(5), 946–958. 10.1111/add.14878

Hughto, J. M., Gordon, L. K., Stopka, T. J., Case, P., Palacios, W. R., Tapper, A., & Green, T. C. (2022). Understanding opioid overdose risk and response preparedness among people who use cocaine and other drugs: Mixed-methods findings from a large, multi-city study. Substance Abuse, 43(1), 465–478. 10.1080/08897077.2021.1946893

Imai, K., Jo, B., & Stuart, E. A. (2011). Commentary: Using potential outcomes to understand causal mediation analysis. Multivariate Behavioral Research, 46(5), 861–873. 10.1080/00273171.2011.606743

Jacoby, J. L., Crowley, L. M., Cannon, R. D., Weaver, K. D., Henry-Morrow, T. K., Henry, K. A., Kayne, A. N., Urban, C. E., Gyory, R. A., & McCarthy, J. F. (2020). Pennsylvania law enforcement use of Narcan. The American journal of Emergency Medicine, 38(9), 1944–1946. 10.1016/j.ajem.2020.01.051

Jones, A. A., Shearer, R. D., Segel, J. E., Santos-Lozada, A., Strong-Jones, S., Vest, N., Texeira da Silva, D., Khatri, U. G., & Winkelman, T. N. A. (2023a). Opioid and stimulant attributed treatment admissions and fatal overdoses: Using national surveillance data to examine the intersection of race, sex, and polysubstance use, 1992–2020. Drug and Alcohol Dependence, 249, 109946. 10.1016/j.drugalcdep.2023.109946

Jones, A., Santos-Lozada, A., Perez-Brumer, A., Latkin, C., Shoptaw, S., & El-Bassel, N. (2023b). Age-specific disparities in fatal drug overdoses highest among older black adults and American Indian/Alaska native individuals of all ages in the United States, 2015-2020. International Journal of Drug Policy, 114, 103977. 10.1016/j.drugpo.2023.103977

Jones, A. A., Schneider, K. E., Tobin, K. E., O’Sullivan, D., & Latkin, C. A. (2023c). Daily opioid and stimulant co-use and nonfatal overdoses in the context of social disadvantage: findings on marginalized populations. Journal of Substance Use and Addiction Treatment, 151, 208986. 10.1016/j.josat.2023.208986

Jones, C. M., Bekheet, F., Park, J. N., & Alexander, G. C. (2020). The evolving overdose epidemic: Synthetic opioids and rising stimulant-related harms. Epidemiologic Reviews, 42(1), 154–166. doi: 10.1093/epirev/mxaa011

Jones, C. M., Warner, M., Hedegaard, H., & Compton, W. (2019). Data quality considerations when using county-level opioid overdose death rates to inform policy and practice. Drug and Alcohol Dependence, 204, 107549. doi: 10.1016/j.drugalcdep.2019.107549

Joudrey, P. J., Khan, M. R., Wang, E. A., Scheidell, J. D., Edelman, E. J., McInnes, D. K., & Fox, A. D. (2019). A conceptual model for understanding post-release opioid-related overdose risk. Addiction Science & Clinical Practice, 14, 1–14. 10.1186/s13722-019-0145-5

JTIC. (2018). Pennsylvania system tracks and centralizes drug overdose information. TechBeat. https://www.ojp.gov/pdffiles1/nij/nlectc/251793.pdf

Karamouzian, M., Cui, Z., Hayashi, K., Debeck, K., Reddon, H., Buxton, J. A., & Kerr, T. (2024). Longitudinal polysubstance use patterns and non-fatal overdose: A repeated measures latent class analysis. International Journal of Drug Policy, 104301. 10.1016/j.drugpo.2023.104301

King, B., Patel, R., & Rishworth, A. (2021). Assessing the relationships between COVID-19 stay-at-home orders and opioid overdoses in the State of Pennsylvania. Journal of Drug Issues, 51(4), 648–660. 10.1177/00220426211006362

Krawczyk, N., Eisenberg, M., Schneider, K. E., Richards, T. M., Lyons, B. C., Jackson, K., … & Saloner, B. (2020). Predictors of overdose death among high-risk emergency department patients with substance-related encounters: a data linkage cohort study. Annals of Emergency Medicine, 75(1), 1–12. doi: 10.1016/j.annemergmed.2019.07.014

Korona-Bailey, J. A., Nechuta, S., Golladay, M., Moses, J., Bastasch, O., & Krishnaswami, S. (2021). Characteristics of fatal opioid overdoses with stimulant involvement in Tennessee: A descriptive study using 2018 State Unintentional Drug Overdose Reporting System Data. Annals of Epidemiology, 58, 149–155. 10.1016/j.annepidem.2021.03.004

Korthuis, P. T., Cook, R. R., Foot, C. A., Leichtling, G., Tsui, J. I., Stopka, T. J., Leahy, J., Jenkins, W. D., Baker, R., Chan, B., Crane, H. M., Cooper, H. L., Feinberg, J., Zule, W. A., Go, V. F., Estadt, A. T., Nance, R. M., Smith, G. S., Westergaard, R. P., Van Ham, B., Brown, R., & Young, A. M. (2022). Association of methamphetamine and opioid use with nonfatal overdose in rural communities. JAMA Network Open, 5(8), e2226544–e2226544. doi:10.1001/jamanetworkopen.2022.26544

Lieberman, A., & Davis. C. (2023). State non-fatal overdose reporting requirements: Fact sheet. The Network for Public Health Law. https://www.networkforphl.org/resources/state-non-fatal-overdose-reporting-requirements/

MacKinnon, D. P., Valente, M. J., & Gonzalez, O. (2020). The correspondence between causal and traditional mediation analysis: The link is the mediator by treatment interaction. Prevention Science, 21, 147–157. 10.1007/s11121-019-01076-4

Mitchell, O., & Caudy, M. S. (2015). Examining racial disparities in drug arrests. Justice Quarterly, 32(2), 288–313. 10.1080/07418825.2012.761721

Morris, N. P., & Kleinman, R. A. (2020). Overdose reversals save lives–period. JAMA Psychiatry, 77(4), 339–340. doi:10.1001/jamapsychiatry.2019.4000

Murphy, J., & Russell, B. (2020). Police officers’ views of naloxone and drug treatment: Does greater overdose response lead to more negativity? Journal of Drug Issues, 50(4), 455–471. 10.1177/00220426209213

Palis, H., Xavier, C., Dobrer, S., Desai, R., Sedgemore, K. O., Scow, M., Lock, K., Gan, W., & Slaunwhite, A. (2022). Concurrent use of opioids and stimulants and risk of fatal overdose: A cohort study. BMC Public Health, 22(1), 2084. 10.1186/s12889-022-14506-w

Pennsylvania State Data Center. (2022). 2020 Census Redistricting Data: Racial & Ethnic Diversity in Pennsylvania. https://pasdc.hbg.psu.edu/sdc/pasdc_files/researchbriefs/January_2022.pdf

Pennsylvania State Police. (2024). Overdose Information Network Data CY January 2018-Current Monthly County State Police. https://data.pa.gov/Opioid-Related/Overdose-Information-Network-Data-CY-January-2018-/hbkk-dwy3/about_data

Pennsylvania State Police. (n.d.). What is ODIN? Overdose Information Network. https://www.pavtn.net/media/1lcf20ws/odin-information-flyer.pdf

Pergolizzi Jr, J. V., Varrassi, G., LeQuang, J. A., & Raffa, R. B. (2021). The challenge of polysubstance use overdose. Open Journal of Social Sciences, 9(7), 529–542. doi: 10.4236/jss.2021.97038

Piza, E., Wolff, K., Hatten, D. and Barthuly, B. (2023). Drug overdoses, geographic trajectories, and the influence of built environment and neighborhood characteristics. Health & Place, 79, 102959. 10.1016/j.healthplace.2022.102959

Ray, B., O’Donnell, D., & Kahre, K. (2015). Police officer attitudes towards intranasal naloxone training. Drug and Alcohol Dependence, 146, 107–110. 10.1016/j.drugalcdep.2014.10.026

Ray, B., Lowder, E., Bailey, K., Huynh, P., Benton, R., & Watson, D. (2020). Racial differences in overdose events and polydrug detection in Indianapolis, Indiana. Drug and Alcohol Dependence, 206, 107658. 10.1016/j.drugalcdep.2019.107658

Rhoads, D. (2019). Pennsylvania Overdose Information Network (ODIN): Executive summary. https://www.nascio.org/wp-content/uploads/2020/09/PA-Data-Mgmt-Overdose-Information-Network-NASCIO-2019-FINAL.pdf

Riley, E. D., Hsue, P. Y., & Coffin, P. O. (2022). A chronic condition disguised as an acute event: The case for re-thinking stimulant overdose death. Journal of General Internal Medicine, 37(13), 3462–3464. 10.1007/s11606-022-07692-1

Rzasa Lynn, R., & Galinkin, J. L. (2018). Naloxone dosage for opioid reversal: Current evidence and clinical implications. Therapeutic Advances in Drug Safety, 9(1), 63–88. doi: 10.1177/2042098617744161

S.B. 1152, P.L. 2158. Act No. 158 of 2022. Overdose mapping act-enactment. Pennsylvania General Assembly. https://www.legis.state.pa.us/cfdocs/legis/CH/Public/ucons_pivot_pge.cfm?session=2022&session_ind=0&act_nbr=0158.

Spencer, M. R., Miniño, A. M., & Garnett, M. F. (2023). Co-involvement of opioids in drug overdose deaths involving cocaine and psychostimulants, 2011–2021. NCHS Data Brief, 474. 10.15620/cdc:129733

Segel, J. E., Shearer, R. D., Jones, A. A., Khatri, U. G., Howell, B. A., Crowley, D. M., Sterner, G., Vest, N., Texeira da Silva, D., & Winkelman, T. N. (2024). Understanding regional patterns of overdose deaths related to opioids and psychostimulants. Substance Use & Misuse, 59(4), 558–566. 10.1080/10826084.2023.2287220

Shastry, S., Shulman, J., Aldy, K., Brent, J., Wax, P., Manini, A. F., & Toxicology Investigators Consortium Fentalog Study Group. (2024). Psychostimulant drug co-ingestion in non-fatal opioid overdose. Drug and Alcohol Dependence Reports, 10, 100223. 10.1016/j.dadr.2024.100223

Shufflebarger, E. F., Reynolds, L. M., McNellage, L., Booth, J. S., Brown, J., Edwards, A. R., Li, L., Robinett, D. A., & Walter, L. A. (2024). Fentanyl-positive urine drug screens in the emergency department: Association with intentional opioid misuse and racial disparities. Drug and Alcohol Dependence Reports, 100269. 10.1016/j.dadr.2024.100269

Silverstein, S. M., Daniulaityte, R., Getz, K., & Zule, W. (2021). “It’s crazy what meth can help you do”: Lay beliefs, practices, and experiences of using methamphetamine to self-treat symptoms of opioid withdrawal. Substance Use & Misuse, 56(11), 1687–1696. 10.1080/10826084.2021.1949612

Silverstein, S., Perdue, T., Baluran, D., Kendrick, E., Larsen, K., Mack, N., & Hassan, R. (2024). The role of law enforcement officers in overdose response: Insights from a qualitative study with people who use drugs and law enforcement officers. Drug and Alcohol Dependence, 260, Supplement, 110703. 10.1016/j.drugalcdep.2023.110703

Substance Abuse and Mental Health Services Administration. (2021). Treatment for Stimulant Use Disorders. Chapter 3-Medical Aspects of Stimulant Use Disorders. https://www.ncbi.nlm.nih.gov/books/NBK576550/

StataCorp (2023). Stata 18 Base Reference Manual. Stata Press.

Valente, P. K., Bazzi, A. R., Childs, E., Salhaney, P., Earlywine, J., Olson, J., Biancarelli, D. L., Marshall, B. D. L., & Biello, K. B. (2020). Patterns, contexts, and motivations for polysubstance use among people who inject drugs in non-urban settings in the US Northeast. International Journal of Drug Policy, 85, 102934. 10.1016/j.drugpo.2020.102934

Wagner, K. D., Fiuty, P., Page, K., Tracy, E. C., Nocera, M., Miller, C. W., Tarhuni, L. J., & Dasgupta, N. (2023). Prevalence of fentanyl in methamphetamine and cocaine samples collected by community-based drug checking services. Drug and Alcohol Dependence, 252, 110985.

Wagner, K. D., Bovet, L. J., Haynes, B., Joshua, A., & Davidson, P. J. (2016). Training law enforcement to respond to opioid overdose with naloxone: Impact on knowledge, attitudes, and interactions with community members. Drug and Alcohol Dependence, 165, 22–28. 10.1016/j.drugalcdep.2016.05.008

Watson, D. P., Ray, B., Robison, L., Huynh, P., Sightes, E., Walker, L. S., Brucker, K., & Duwve, J. (2018). Lay responder naloxone access and Good Samaritan law compliance: postcard survey results from 20 Indiana counties. Harm Reduction Journal, 15, 1–8. 10.1186/s12954-018-0226-x

Williams, R. H., & Erickson, T. (2000). Emergency diagnosis of opioid intoxication. Laboratory Medicine, 31(6), 334–342. 10.1309/QY8T-KGBN-BVA6-H706

Wootson, C. R. (2017). Why this Ohio sheriff refuses to let his deputies carry Narcan to reverse overdoses. The Washington Post. https://www.washingtonpost.com/news/to-your-health/wp/2017/07/08/an-ohio-countys-deputies-could-reverse-heroin-overdoses-the-sheriff-wont-let-them/

